# A scoping review of guidelines on reporting and assessing dynamic mathematical models of infectious diseases

**DOI:** 10.1101/2024.11.27.24318060

**Authors:** Madhav Chaturvedi, Antonia Bartz, Claudia M Denkinger, Carolina Klett-Tammen, Mirjam Kretzschmar, Alexander Kuhlmann, Berit Lange, Florian M. Marx, Rafael Mikolajczyk, Ina Monsef, Hoa Thi Nguyen, Janik Suer, Nicole Skoetz, Veronika K Jaeger, André Karch

## Abstract

**Background:** Mathematical models are essential for guiding public health policy decisions to combat the spread of infectious diseases. Nevertheless, there are no widely-used quality assessment tools that can be used to assess the quality of infectious disease modelling studies. There is also a lack of commonly accepted and used reporting guidelines that researchers can follow to improve the comprehensibility, transparency, and credibility of their publication. This scoping review identified common themes in existing reporting and quality assessment guidance for infectious disease modelling studies and adjacent fields of research.

**Methods:** We conducted temporally-unrestricted searches on Medline (via Ovid), Web of Science, medRxiv, and bioRxiv on January 4^th^, 2024 to find articles that provide guidance on writing or assessing modelling studies within infectious disease modelling and adjacent fields like health economics. Articles were double-screened for eligibility via title-and-abstract screening and full-text screening. Recommendations made by eligible articles were classified into 33 subdimensions which were categorised into seven dimensions (applicability; model structure; parameterisation and calibration; validity; uncertainty; interpretation; and reproducibility, clarity, and transparency). We followed the PRISMA extension for reporting scoping reviews.

**Results:** Fifty-two articles were included in our final review. All dimensions except for interpretation were covered by most articles (79%-98%). However, we found substantial heterogeneity in the frequency with which subdimensions were addressed (12%-96%). Subdimensions pertaining to study design, assumptions about model structure, handling of parameter uncertainty, and transparency about parameter values were mentioned in most articles (85%-96%); conversely, discussions about auxiliary details regarding publication, software implementation, parsimony, and predictive validity were covered less frequently (23%-31%).

**Conclusion:** This review reaffirms the lack of commonly used guidelines on reporting and assessing infectious disease models. Furthermore, it identifies common topics and recommendations from neighbouring fields which can inform the development of standardised guidelines for infectious disease modelling.

## Introduction

Infectious diseases remain a leading cause of morbidity and mortality worldwide (1). Nowadays, mathematical modelling is increasingly used to study the dynamics, transmission, and control of infectious diseases. The role of modelling as an indispensable tool for guiding public health policy was exemplified during the COVID-19 pandemic (2). Modelling fields beyond infectious disease modelling have undertaken considerable efforts to standardise reporting and quality assessment of modelling studies (3,4). However, to our knowledge, there remains a lack of widely-used guidelines for the reporting and assessment of infectious disease modelling studies.

Inadequate reporting of research and a lack of standard quality assessment methods are major concerns that can cause misinterpretation and an inability to assess the validity of a study. Reporting guidelines and quality assessment are tools that aim to counteract these shortcomings (5). Reporting guidelines provide structured frameworks to transparently report study methods, results, and interpretations, ensuring that key information is consistently reported (6,7). Quality assessment tools enable a systematic evaluation of studies by identifying potential sources of bias or systematic error (8). Adhering to reporting guidelines allows researchers to enhance the transparency, reliability, and credibility of their research. Conducting quality assessments enables readers, researchers, and policy makers to critically assess the reliability of study results and compare different studies on the same key question to each other.

Having standardised reporting guidelines and quality assessment tools specifically for infectious disease modelling studies would help not only researchers but also public health authorities and decisionmakers to better judge and assess evidence from modelling studies, ultimately contributing to more comprehensive understanding of infection dynamics, transmission, and control strategies.

In this scoping review, we identified existing guidance and best practice recommendations—for reporting or assessing studies—in the field of infectious disease modelling, as well as standardised guidelines that exist in adjacent fields. Specifically, we determined common themes across the identified publications which could be used to create a core item set for a standardised reporting guideline and quality assessment tool specific to infectious disease modelling.

## Methods

### Search strategy and article selection for scoping review

We conducted searches on Medline (via Ovid) (9), Web of Science (10), medRxiv (11), and bioRxiv (12) on the 4^th^ of January, 2024 to find articles that provide guidance on writing or assessing modelling studies. The search was not restricted temporally or by language. Further details about these searches, including the search strings used and the number of hits for each search, are given in Supplementary Material (SM) 1. Although the focus of the review was on infectious disease modelling, we included search terms to find articles on modelling studies in other fields—especially health economics—since some of the fundamental reporting and assessment tools for these types of models may be applicable to infectious disease models as well.

The articles found by these searches were aggregated and, after the removal of duplicates, filtered through double screening. Screening was conducted in two rounds—first a title-and-abstract screening (AB, MC, VKJ), and then a full-text screening (AB, MC, JS). Conflicts at both screening stages were resolved by discussion (AB, MC, JS, VKJ). When articles were reviews in themselves, the individual articles included in the review were also considered for inclusion, and the review article was only included if the authors of the review made suggestions or recommendations of their own.

Inclusion and exclusion criteria for the screening rounds are given in SM1. A full protocol for the scoping review is available on the Open Science Framework platform (13).

### Identification of articles via other methods

In an effort to include articles beyond the scope of our search strategy, we considered additional methods of article identification. We included guideline articles known to us or experts in the field with whom we communicated that had not appeared in the search, as well as relevant articles found by snowballing. Additionally, we searched the EQUATOR (Enhancing the QUAlity and Transparency Of health Research) Network library (14) to determine whether any reporting guidelines for infectious disease modelling studies had been published there or had been reported to be under development.

Furthermore, we looked at recently published infectious disease modelling studies in order to determine whether reporting guidelines are mentioned in any of these publications. We went through 100 recent modelling publications to establish if any of the authors mentioned following a reporting guideline (AB, VKJ). These 100 publications were identified by single-screening the title and abstract of a random selection of infectious disease modelling publications published between 1^st^ January 2019 and 22^nd^ January 2024 (MC). Further details about this search are provided in SM1. If deemed relevant, any guidelines identified via this strategy were included in the list of articles to be extracted.

### Data Extraction and Analysis

Recommendations made in the included articles were extracted in a Microsoft Excel spreadsheet (SM2). The initial layout of the spreadsheet was determined by combining and classifying the points mentioned by two quality assessment tools (15,16)—which we had previously used to assess the quality of several modelling studies—into broad dimensions and subdimensions (AB, MC, VKJ, AKa). This initial structure provided an overview of dimensions and subdimensions we expected additional guidelines to cover, thereby acting as a foundation for the extraction of all other identified guidelines.

For each article, we single-extracted relevant (i.e. applicable to dynamic infectious disease models) information from provided checklists, headings, or text and categorised them into the existing subdimensions or added new subdimensions when necessary (AB, MC, VKJ, Aka). For validation purposes, twenty of the articles were double-extracted.

All figures in the manuscript were created using the ggplot2 package in R (17). We adhered to the PRISMA checklist extension for scoping reviews (18) while writing the manuscript (SM4).

## Results

Details about the number of articles that progressed through each round of screening can be found in Figure 1. We identified 8182 articles from the queried databases. After title-and-abstract and full-text screening, we were left with 45 articles. We supplemented these with six articles known to us that had not appeared in our search (19–24). Our perusal of 100 random infectious disease modelling studies published in the past five years revealed that only one study mentioned using a reporting guideline (25). This identified guideline (26) was added to the list of articles eligible for extraction leading to a total of 52 articles eligible for extraction (27–30)(31–34)(35,36)(37,38)(39)(40)(41)(42–44). Of these, 11 (21%) were reviews of literature themselves (3,23,29,45–52), but added their own recommendations based on their findings.

**Figure 1.**
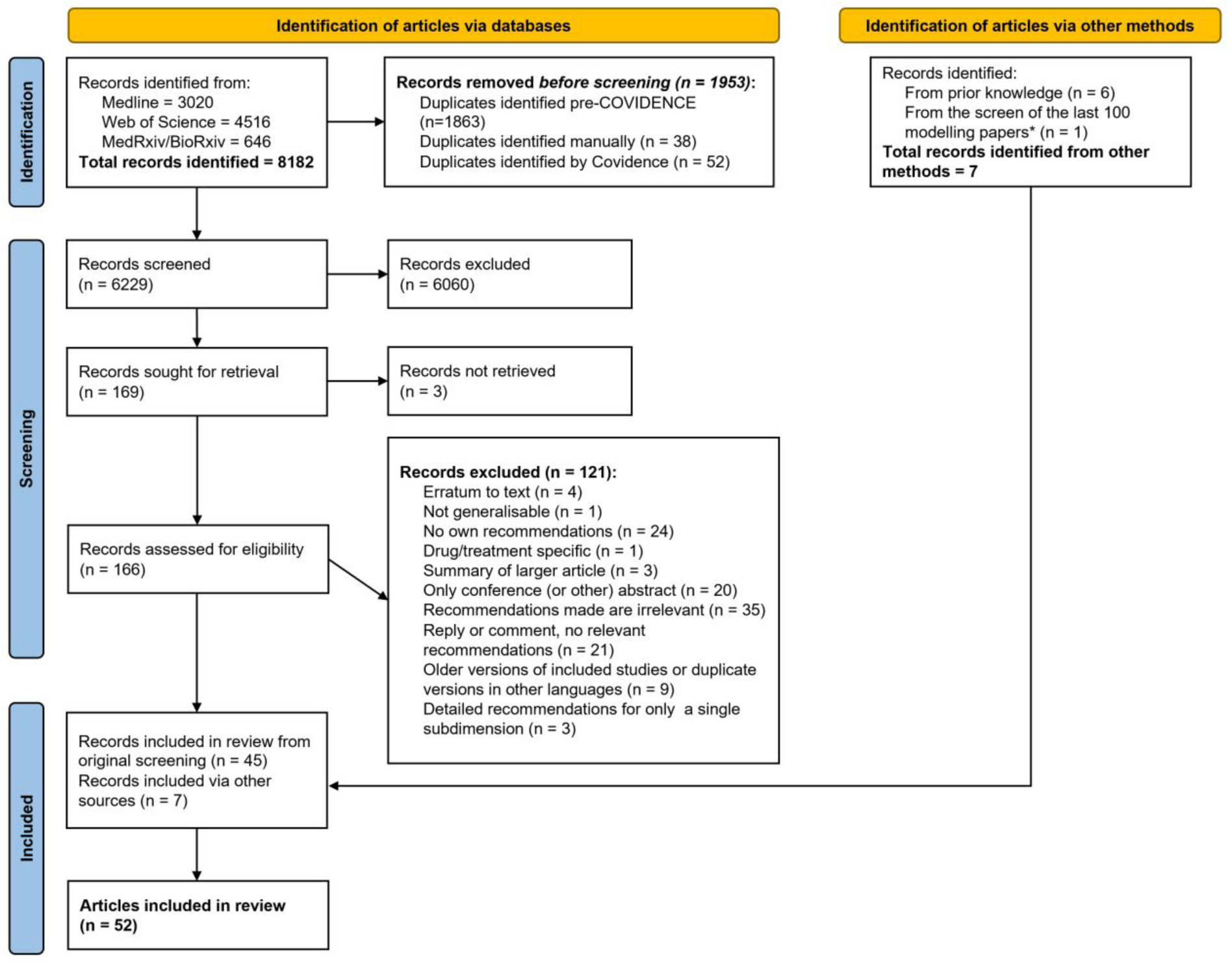
PRISMA flowchart of articles included in the scoping review.

### Purpose

The identified articles included best practice model development guidelines, reporting recommendations, and quality assessment tools, with some articles serving several purposes. Twenty-two of the identified articles (42%) explicitly mentioned that the presented guideline, recommendation, or tool had undergone a consensus-building process during development (23,27,28,30,39,41,49,51,53–66). Seventeen of the 52 articles (33%) (2,20,23,25,27,44,51,52,57,62,64–69) gave recommendations, checklists, or guidance on how to report models and modelling studies in an article, ten articles (19%) gave recommendations for developing models and conducting modelling studies (19,22,27,30,41,49,51,54,66,73), and 19 articles (37%) described tools or recommendations for assessing the quality of existing modelling studies (20,24,45,46,48,50,52,53,56,57,60,61,74–79) (including one article specifically about multi-model comparisons (56)). Furthermore, ten articles 19%) presented best practice or good practice recommendations, which usually included aspects of both model development and reporting (29,39,58,59,62–64,80–82); these recommendations can easily be used to assess the quality of studies as well.

### Topic

Of the 52 articles, 33 (62%) were concerned with models or modelling studies for health economic evaluations (22,23,27–30,41,46–52,54,57–61,63–68,70,74–77,80,82). Ten of the 52 articles (19%) were about infectious disease models, but they were all narrow in their scope. Two of these 10 focused specifically on modelling drug-resistant infections in healthcare or long-term care settings (72,78), and a further two discussed models specifically designed for epidemic forecasting and analysing emerging outbreaks (55,73). One restricted its scope to individual-based models of HIV transmission (83), and one documented development and reporting guidelines for infectious disease modelling analyses about vaccination interventions to be presented to the German Standing Committee on Vaccination (STIKO) (84). Of the four remaining articles about infectious disease modelling, one presented guidelines for multi-model comparisons of infectious disease interventions (56), one was a review of literature mostly consisting of articles about health economics (85), one provided guidance on how to implement models transparently and reproducibly (71), and one provided a guide to policy-makers for assessing the utility of models for a specific public health question (79).

Seven of the 52 articles (13%) considered a wider scope of models used to answer healthcare questions without being restricted to the health economic perspective or only limited to infectious disease models (3,20,39,53,62,81). There were two articles not explicitly focused on healthcare: one described a protocol for reporting agent-based models (26) and the other gave recommendations for describing simulation models in general regardless of type or purpose (21).

### Dimensions of content

We classified recommendations from the selected articles into seven broad dimensions, each representing a different facet of a modelling study. These dimensions were: applicability; model structure; parameterisation and calibration; validity; uncertainty; interpretation; and reproducibility, clarity, and transparency (Figure 2). Where appropriate, we further divided these dimensions into subdimensions; these are summarised in Table 1 and Figure 3.

**Figure 2.**
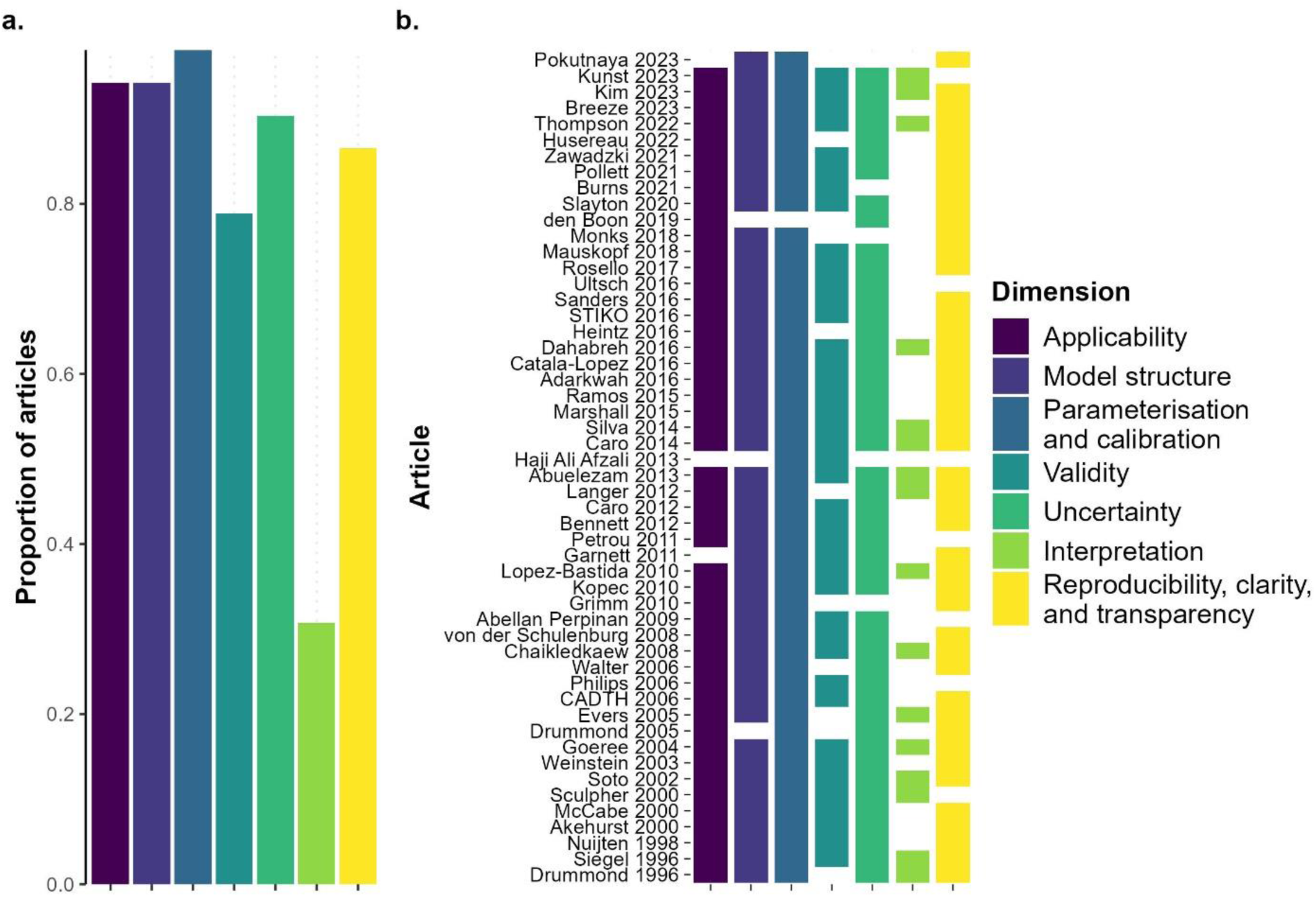
Prevalence of each dimension in the reviewed articles. (a) Overall proportion of articles mentioning each dimension. (b) Presence or absence of each dimension split up by article. The data used for these plots can be found in SM2.

**Figure 3.**
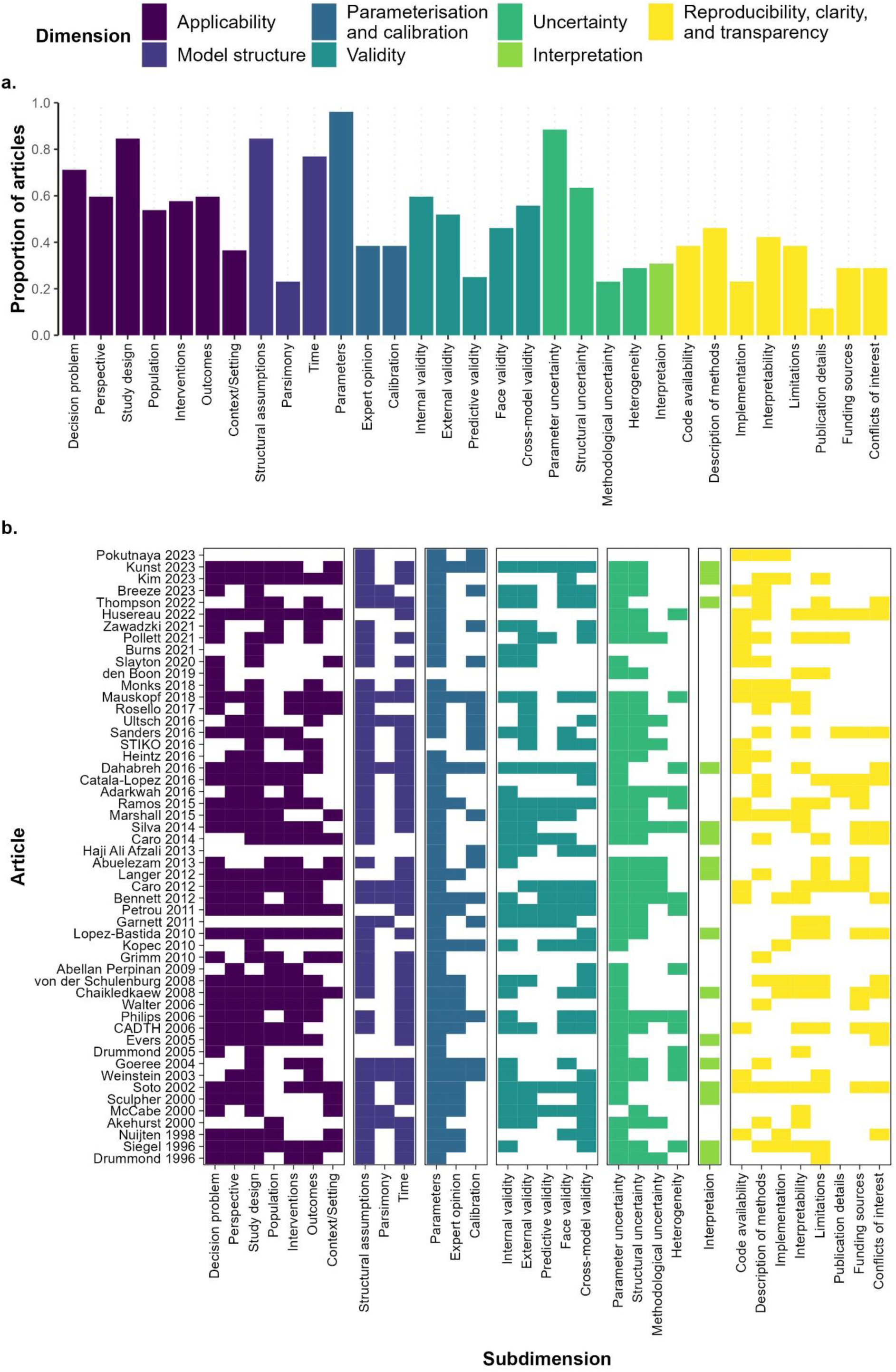
Prevalence of each subdimension in the reviewed articles. (a) Overall proportion of articles mentioning each subdimension. (b) Presence or absence of each subdimension split up by article. The data used for these plots can be found in SM2.

**Table 1.** Overview of dimensions and subdimensions as well as a brief summarised question that addresses the elements mentioned by articles (for reporting as well as for quality assessment) that were extracted and classified into each subdimension.

| Dimension/Subdimension | Assessment/Reporting Criteria |
| --- | --- |
| **1. Applicability** |  |
| 1.1 Decision problem | Is there a clear statement of the aim/decision problem? |
| 1.2 Perspective | Is the perspective (i.e. scope (e.g. societal or a specific economics/healthcare system) considered in the model) of the study clearly stated and/or relevant? |
| 1.3 Study design | Is the study design clearly stated and/or applicable to the decision problem? |
| 1.4 Population | Is the population clearly stated and/or applicable to the decision problem? |
| 1.5 Interventions | Is there a comprehensive consideration of the interventions and/or comparators? |
| 1.6 Outcomes | Are the outcomes clearly stated and/or applicable to the decision problem? |
| 1.7 Context/Setting | Is the context/setting clearly stated and/or applicable to the decision problem? |
| **2. Model structure** |  |
| 2.1 Structural assumptions | Are structural and methodological assumptions clearly stated and reasonable? |
| 2.2 Parsimony | Is the model as simple as possible but as complex as necessary? |
| 2.3 Time | Is the time horizon and/or time step clearly stated and appropriate? |
| **3. Parameterisation and Calibration** |  |
| 3.1 Parameters | Are parameter values transparent and reasonable?<br>Are data sources and their translation into parameter values transparent and justified? |
| 3.2 Expert opinion | If applicable, has the methodology used to elicit expert opinion for parameterisation been sufficiently described? |
| 3.3 Calibration | If applicable, were methods and data sources used for calibration described in sufficient detail? Were the subsequent results presented clearly? |
| **4. Validity** |  |
| 4.1 Internal validity | Has evidence of the mathematical soundness and correctness of model implementation been given? |
| 4.2 External validity | Has the model been validated against independent data sources? |
| 4.3 Predictive validity | Has the ability of the model to predict future events been shown? |
| 4.4 Face validity | Is the model structure plausible? Have any counterintuitive results been explained? |
| 4.5 Cross-model validity | Has the model been validated against similar models? |
| **5. Uncertainty** |  |
| 5.1 Parameter uncertainty | Have the methods used to assess uncertainty surrounding parameters been clearly described? |
| 5.2 Structural uncertainty | Have the methods used to assess uncertainty arising from structural assumptions been clearly described? |
| 5.3 Methodological uncertainty | Have the methods used to assess uncertainty stemming from the choice of methodology been clearly described? |
| 5.4 Heterogeneity | Have the methods used to assess variability in results across subgroups been clearly described? |
| **6. Interpretation** |  |
| 6.1 Interpretation | Is the presented interpretation of the results reasonable, fair, and/or balanced? |
| **7. Reproducibility, clarity, and transparency** |  |
| 7.1 Code availability | Are the code/model/data available? |
| 7.2 Description of methods | Are methods described transparently and in enough detail that they can be reproduced? |
| 7.3 Implementation | Has the choice of software been stated and justified? |
| 7.4 Interpretability | Is there sufficient non-technical documentation and description? |
| 7.5 Limitations | Are limitations clearly described? |
| 7.6 Publication details | Is the type of study clearly identified in the title or abstract? Has the model developer been stated? |
| 7.7 Funding sources | Are the funding sources and the role of the funder in the study clearly stated? |
| 7.8 Conflicts of interest | Are there any potential conflicts of interest? |

Several similarities emerged when comparing the dimensions and subdimensions covered in the different articles. All dimensions except for interpretation were covered by at least 79% of articles, with applicability being covered in all but two articles, and parameterisation and calibration in all but one. Unsurprisingly, there was more variation when focusing on subdimensions, with prevalence of the different subdimensions ranging from 12% (publication details) to 96% (parameter transparency). A detailed breakdown of the dimensions and subdimensions, including the frequency with which they were mentioned, can be found below.

### 1. Applicability

This dimension includes checklist items that are concerned with the aim or decision problem for a modelling study, and the applicability of individual elements of the modelling study to said aim or decision problem. All but three of the 52 analysed articles (94%) addressed this dimension, in one or more subdimensions (Figure 2, Figure 3). Of the three that did not, two addressed only specific aspects of reporting models: one focused specifically on the transparent and reproducible implementation of models (71), and one provided recommendations on how to report model validation procedures (47).

#### 1.1. Decision problem

In all, 37 articles (71%) mentioned that the decision problem that a modelling study aims to answer should be clearly stated.

#### 1.1. Perspective

Thirty-one articles (60%) mentioned that the perspective (e.g. societal or a specific health economics/healthcare system) chosen by a study should be clearly stated. This was an especially common subdimension mentioned in health economics articles, with 27 of the 33 articles (82%) pertaining to health economics mentioning it.

#### 1.1. Study design

This subdimension, included in 44 of the 52 articles (85%), considers whether the design of a modelling study is described clearly and whether the appropriateness of the chosen design to achieve the study’s aims is justified. This includes the type of model chosen, the scope of the model, and the analytical framework of the study, among other things.

#### 1.1. Population

Of the 52 articles, 28 (54%) mentioned that the population considered in the modelling study (including all relevant demographic characteristics) should be clearly described and should be relevant to the decision problem of the study.

#### 1.1. Interventions

Thirty articles (58%) stated that modelling studies should consider all relevant interventions or justify any exclusions.

#### 1.1. Outcomes

Thirty-one articles (60%) touched upon the outcomes considered in a study, asking modellers to properly describe the outcomes considered or instructing readers to assess whether the outcomes considered were relevant to the stated decision problem and whether any relevant outcomes had been omitted.

#### 1.1. Context/Setting

Nineteen articles (37%) mentioned that the setting (geographical, environmental, etc.) and context (economic, political, infrastructural, etc.) of the decision problem should be well-described and well-represented in a modelling study.

### 2. Model structure

This dimension is concerned with the description and justification of the model structure, including any underlying assumptions, chosen by the researchers conducting a modelling study. Overall, 49 of the 52 analysed articles (94%) suggested topics that could be categorised into one or more subdimensions of model structure (Figure 2, Figure 3).

#### 2.1. Structural Assumptions

A total of 44 articles (85%) addressed the structural assumptions behind a model in a modelling study. Articles providing guidance on reporting mentioned that all structural assumptions should be explicitly and clearly reported, and articles providing guidance for quality assessment instructed the reader to assess whether structural assumptions were justified and consistent with existing knowledge about the phenomenon being modelled. Nine out of the 10 (90%) articles concerned with infectious disease modelling addressed this subdimension.

#### 2.2. Parsimony

Twelve articles (23%) mentioned that the model used in a modelling study should be as simple as possible but complex enough to incorporate all relevant interactions, with any simplifications being justified.

#### 2.3. Time

Overall, 40 articles (77%) explicitly considered how different timeframes were considered and reported in a modelling study, and whether the choices were justified; this included the time horizon or study period the model was run for, as well as the cycle length or time step considered in the model.

### 3. Parameterisation and calibration

This dimension comprises considerations of the parameterisation of the model(s) in a modelling study, both directly from existing literature or data, or indirectly via calibration/model fitting. All analysed articles except the one that focused on multi-model comparisons rather than single modelling studies (56) mentioned model parameterisation (98%), classifiable into the following subdimensions (Figure 2, Figure 3).

#### 3.1. Parameters

All but two of the analysed articles (96%) included discussions of parameter values obtained from existing data, either directly from the literature or after transformation of data. This included the reporting of parameter values (including their sources or a justification), data sources, and data handling and transformation, and whether the parameter values obtained this way were credible.

#### 3.2. Expert opinion

Twenty articles (38%) explicitly discussed the use of expert opinion during model parameterisation, in particular with respect to the reporting of the methodology used to elicit such opinions.

#### 3.3. Calibration

Twenty articles (38%) addressed the calibration of a model for the purposes of parameterisation; this was done by focusing on the sources of data used for calibration, the methodology chosen, and the presentation and discussion of the outcomes. Calibration was an especially common topic in articles specific to infectious disease modelling studies, with seven of the 10 such articles (70%) making recommendations for the reporting or assessment of model calibration.

### 4. Validity

This subdimension comprises suggestions about the reporting or quality of model validity and the validation process employed in a study. The subdimensions correspond to the different types of model validity—internal, external, predictive, face, and cross-model. Of the 52 extracted articles, 41 (79%) provided guidance for at least one of the different types of validity (Figure 2, Figure 3).

#### 4.1. Internal validity

Discussion about evidence and reporting of internal validity of the model(s) used in a study, i.e. the soundness of the mathematical logic of the model, the absence of bugs in the code used to implement the model, etc., was present in 31 articles (60%).

#### 4.2. External validity

Guidance about external validation, i.e. the validation of a model against independent data sets not used for model calibration, were included in 27 of the 52 articles (52%). Suggestions were sometimes contradictory across articles. For example, some articles mentioned that some data can be withheld from calibration to instead be used for validation (15,84), while others recommended that all data should be used for parameterisation and calibration rather than being held back for validation (29).

#### 4.3. Predictive validity

Predictive validity, arguably a type of external validity, is the ability of a model to accurately predict future occurrences. Thirteen articles (25%) made suggestions about the evaluation of predictive validity, while simultaneously conceding that this is not always possible.

#### 4.4. Face validity

Face validity of a modelling study was mentioned by 24 articles (46%). Articles about reporting tended to emphasize the explanation of any counterintuitive results and the presentation of outcomes of expert assessments of face validity and plausibility. Articles providing guidance on quality assessment often instructed the reader to assess the face validity of the study for themselves.

#### 4.5. Cross-model validity

A total of 29 articles (56%) discussed validation of a model against other models or modelling studies, covering the identification and justification of the models used for comparison, the methodology used, and a discussion of the results, especially regarding any discrepancies or differences between the model results.

### 5. Uncertainty

This dimension comprises guidance about the handling of uncertainty in modelling studies, including assessments of the sensitivity of the results to this uncertainty. The subdimensions correspond to the different types of uncertainty present in such a study. All but five of the analysed articles (90%) contained items about handling one or more of the following types of uncertainty.

#### 5.1. Parameter uncertainty

Forty-six articles (88%) discussed parameter uncertainty, i.e. uncertainty due to imprecise parameter estimates. These articles touched on the reporting of parameter uncertainty, sensitivity analysis methods to assess parameter uncertainty, and adequate discussion of the sensitivity analyses and uncertainty around results due to parameter uncertainty.

#### 5.2. Structural uncertainty

Assessment of uncertainty caused by structural assumptions, for instance by examining alternative model structures, was discussed in 33 articles (63%).

#### 5.3. Methodological uncertainty

Twelve articles (23%) mentioned the assessment of uncertainty caused by methodological choices and assumptions made by the researchers in a modelling study.

#### 5.4. Heterogeneity

Fifteen articles (29%), 14 of which were concerned with health economic modelling evaluations, mentioned the assessment of uncertainty caused by heterogeneity in the study population and variability of results across different subgroups (e.g. different age groups or geographical regions) in the target population.

### 6. Interpretation

This dimension, consisting of a singular subdimension, is concerned with statements—present in 16 articles (31%)—about whether the authors’ interpretation of the results of a modelling study are unbiased, reasonable, substantiated by the results, and presented with a discussion of the limitations and caveats (Figure 2, Figure 3).

### 7. Reproducibility, clarity, and transparency

This dimension contains points of discussion and guidance pertaining to reproducibility of a modelling study, interpretability of the results by the target audience and laypeople, and transparency with respect to methods, funding, and the limitations of the study. Of the 52 articles extracted, 45 (87%) had statements pertaining to topics under this dimension (Figure 2, Figure 3).

#### 7.1. Code availability

Twenty articles (38%) touched upon public availability of code and technical documentation, mentioning that the model should be publicly available or made available upon request. If the code cannot be made available, a reason for this should be given.

#### 7.2. Description of methods

In total, 24 articles (46%) mentioned that the methods of a modelling study should be described transparently and in enough detail to be reproducible.

#### 7.3. Implementation

Twelve articles (23%) stated that the software and programming languages used during a modelling study should be clearly stated, with two articles stating that the choice of software should be justified.

#### 7.4. Interpretability

A total of 22 articles (42%) mentioned that the reporting of methods and results in a modelling study should be done in such a way as to be accessible to the target audience and any interested reader (especially policy makers) regardless of their technical background. This includes using non-technical language and documentation. At a minimum, a non-technical summary and conclusion should be available.

#### 7.5. Limitations

Twenty of the 52 articles (38%) suggested the need for transparent discussion of the limitations (and strengths) of the modelling approach and results of a study.

#### 7.6. Publication details

Six articles (12%) suggested auxiliary details that should be included in the publication of a modelling study, including disclosure of author contributions and an identification of the type of study in the title or abstract.

#### 7.7. Funding sources

Fifteen articles (29%) mentioned that the funders of a modelling study and their role in the study should be clearly described.

#### 7.8. Conflicts of interest

Fifteen articles (29%) advised the clear disclosure of any conflicts of interest and steps taken to mitigate these conflicts.

## Discussion

With this study, we aimed to assess whether any commonly used reporting guidelines or quality assessment tools exist for infectious disease modelling, and to identify common dimensions and subdimensions appearing in modelling studies from neighbouring fields of research. Although no commonly used infectious disease-specific modelling guidelines were identified, we found 52 relevant articles from which a plethora of common dimensions and subdimensions could be identified.

The transparent reporting of parameter values and their sources was the most frequently mentioned subdimension across all studies. Having a clear statement of the research question or decision problem for a modelling study and a well-described study design that is appropriate for the decision problem were also recommended by the majority of articles. A clear description and justification of the structural assumptions as well as the assessment of uncertainty around parameters were also prominent subdimensions. Ensuring reproducibility of studies was also a common recommendation, manifesting most often as recommending the sharing of data, code, and models, and ensuring a thorough description of methods. These subdimensions could form a minimal set of core items for inclusion in a key guideline for infectious disease modelling. This set is however by no means exhaustive. Many subdimensions appearing less frequently are equally important; this includes, for example, those touching on reporting conflicts of interest, the validation of models before use, or the interpretability of the study by a non-technical audience (especially policy makers).

However, the subdimensions have varying applicability and relevance to infectious disease modelling. For example, structural assumptions and parameter values are crucial to an infectious disease model—as with any other model—and the corresponding subdimensions were addressed by a majority of the articles concerned with infectious disease modelling. On the other hand, establishing predictive validity may be impossible in some situations where infectious disease models are needed, and a choice of perspective is much more relevant to health economic evaluations than infectious disease models. Thus, when applying the findings of these studies to recommendations for infectious disease models, it is important to keep the diversity of topics covered in this review in mind.

The definitions of the various subdimensions in our study are subjective, as are the classifications of recommendations into these subdimensions. This means that some subdimensions are broader than others, and that elements covered in individual articles could be classified into several different subdimensions. This would impact the frequency with which certain subdimensions are mentioned. Consequently, the results are, to some extent, an oversimplification of the true complexity and diversity of the articles used in this study. A deeper look at the content of individual articles may be helpful to better understand the depth and intricacies of existing guidelines and quality assessment tools. Nonetheless, our results provide an overview of the numerous dimensions and subdimensions that commonly occur in reporting guidelines and quality assessment tools for modelling studies.

## Conclusion

This review demonstrates the existence of many common topics and recommendations towards standardized reporting guidelines. The more common use of modelling in the context of COVID-19 pandemic supports the need of defining such guidelines. To this end, the common recommendations we have identified in this review of guidelines from neighbouring fields could be adapted to create guidelines for infectious disease modelling. Establishing such recommendations would be monumental in assisting researchers and policy makers by facilitating the inclusion of modelling studies in the public health decision-making process. Therefore, as a follow-up to this review, our next steps will be to develop a quality assessment tool and a reporting guideline by building upon the subdimensions identified in this review.

## Data Availability

The search strategy is available in Supplementary Material (SM) 1. The complete dataset is available in SM2. Code used for creating figures is available in SM3. The PRISMA-ScR Checklist can be found in SM4.

## Statements

### Ethics

N/A

### Author contributions

Conceptualization: MC, AB, VKJ, Aka

Data curation: MC, AB, IM

Formal analysis: MC, AB

Funding acquisition: AKu, BL, RM, VKJ, Aka

Investigation: MC, AB, VKJ, Aka

Interpretation: all authors

Methodology: all authors

Project administration: MC, AB, VKJ, Aka

Resources: AKa

Software: MC, AB, VKJ

Supervision: VKJ, Aka

Validation: VKJ, Aka

Search: MC, AB, IM, VKJ

Screening: MC, AB, JS, VKJ

Visualization: AB, VKJ

Writing - original draft: MC, AB, VKJ, Aka

Writing - review & editing: all authors

