## Supplementary File 1 for "A scoping review of guidelines on reporting and assessing dynamic mathematical models of infectious diseases"

**Guidelines on reporting and assessing mathematical models for infectious disease dynamics: A scoping review**

**Supplement 1. Search strategy**

Inhalt

### **1. Literature search of guidelines and recommendations**

#### 1.1. Search results

| **Date of search for all databases: 04.01.2024** | |
| --- | --- |
| **Database/Register** | **Hits** |
| **MEDLINE ALL (via Ovid)** 1946 to present | **3020** |
| **Web of Science (SCI-expanded, ESCI)** | **4516** |
| **MedRxiv BioRxiv** | **646** |
| **Total** | **8182** |
| **Total (after deduplication)** | **6319** |

#### 1.2. Search strategies

##### **1.2.1 MEDLINE** (via OVID) 1946 to January 03, 2024

\#	Searches

1	exp Guidelines as topic/ or exp Guideline/

2	(guideline* or guidance or (practice adj2 (guide*1 or recommend* or standard*)) or ((good* or best*) and practi*)).ti.

3	(framework* or checklist* or recommend* or concept* or standard* or position paper or reporting or reported or report?).ti.

4	or/1-3

5	((infecti* disease* or communicable disease* or transmission*) adj4 model*).tw.

6	(dynamic adj4 model*).ti.

7	(((economic* or cost?) adj2 evaluation*) or (cost? adj2 effectiveness)).ti.

8	((decision? adj1 analy*) and model*).tw.

9	(model* adj8 stud*).ti.

10	or/5-9

11	4 and 10

##### 1.2.2. Web of Science (Science Citation Expanded 1945 to 2024, Emerging Sources Citation Index 2019 to 2024)

#1	(((TI=(guideline* )) OR TI=(guidance*)) OR TI=( (practice NEAR/2 (guide* or

recommend* or standard*)))) OR TI=(((good* or best*) and practi*))

#2	TI=((framework* or checklist* OR recommend* OR concept* OR standard* OR "position

paper" OR reporting OR reported OR report OR reports))

#3	#2 OR #1

#4	(TI=(("infecti* disease*" NEAR/4 model*) OR (communicable disease NEAR/4 model*)

OR (transmission NEAR/4 model*))) OR AB=(("infecti* disease*" NEAR/4 model*) OR

(communicable disease NEAR/4 model*) OR (transmission NEAR/4 model*))

#5	TI=((dynamic NEAR/4 model*))

#6	TI=(((economic* OR cost OR costs) NEAR/2 evaluation*) OR (cost NEAR/2

effectiveness) OR (costs NEAR/2 effectiveness))

#7	(TI=(((decision? NEAR/1 analy*) AND model*))) OR AB=(((decision? NEAR/1 analy*)

AND model*))

#8	TI=((model* NEAR/8 stud*))

#9	#8 OR #7 OR #6 OR #5 OR #4

#10	#9 AND #3

##### 1.2.3 MedRxiv (<https://www.medrxiv.org/search>)

Advanced search in medRxiv or bioRxiv

Separate searches in title or abstract

"infectious diseases" model* guideline*"; "communicable disease" model* guideline*"; "communicable diseases" model* guideline*"; "transmission* model* guideline*"; decision* model* guideline*; "economic evaluation" guideline*; "economic evaluations" guideline*; "cost evaluation guideline*"; "cost evaluations" guideline*"; "simulation model* guideline*"; "multi modal" comparison* guideline*"; "multimodal" comparison* guideline*"; "infectious diseases" framework* model*"; "communicable disease" model* framework*; "communicable diseases" model* framework*;

Separate searches in title:

"modelling guideline*"; "modeling guideline*"; "modelling guidance*"; "modeling guidance*";modeling* framework*; modelling* framework*; modeling* checklist*; modelling* checklist*; modelling* recommend*; modeling* recommend*; modelling* concept*; modeling* concept*; modelling "position paper"; modeling"position paper"; modelling report*; modelling report*; "infectious disease" model* guidance*; "infectious diseases" model* guidance*; "communicable disease" model* guidance*; "communicable diseases" model* guidance*; transmission* model* guidance*; decision* model* guidance*; "economic evaluation" guidance*; "economic evaluations" guidance*; cost evaluation guidance*; "cost evaluations" guidance*; simulation model* guidance*; "multi modal" comparison* guidance*; "multimodal" comparison* guidance*; "infectious disease" framework*; "transmission* framework*"; "decision* analytic* framework*"; ""economic evaluation" framework*"; ""economic evaluations" framework*"; ""cost evaluation" framework*"; ""cost evaluations" framework*"; ""multi modal" comparison* framework*"; ""multimodal" comparison* framework*"; ""infectious disease" model* checklist*"; ""infectious disease" checklist*"; ""communicable disease" checklist*"; ""communicable diseases" checklist*"; "transmission checklist*"; "decision model* checklist*"; "decision analytic* checklist*"; ""economic evaluation" checklist*"; ""economic evaluations" checklist*"; ""cost evaluation" checklist*"; ""cost evaluations" checklist*"; ""simulation model" checklist*"; ""multi modal comparison" checklist*"; ""multimodal comparison" checklist*"; ""infectious disease" recommend*"; ""infectious diseases" recommend*"; ""communicable disease" recommend*" ; ""communicable diseases" recommend*" ; "transmission model* recommend*"; "decision* model* recommend*"; "economic evaluation" recommend*; "economic evaluations" recommend*; ""cost evaluation" recommend*"; ""cost evaluations" recommend*"; "simulation model* recommend*"; ""multi modal comparison" recommend*"; ""multimodal comparison" recommend*"; ""infectious disease" concept*"; ""infectious diseases" concept*"; ""communicable disease" concept*"; ""communicable diseases" concept*"; "transmission model* concept*"; decision model* concept*; ""economic evaluation" concept*"; ""economic evaluations" concept*"; ""cost evaluation" concept*"; ""cost evaluations" concept*"; "multi modal comparison* concept*"; "multimodal comparison* concept*"; "infectious disease* standard*"; "communicable disease* standard*"; "transmission model* standard*"; "decision model* standard*"; "economic evaluation* standard*"; "simulation model* standard*"; "multi modal comparison* standard*";"multimodal comparison* standard*"; "position paper"; "disease* report*"; "transmission model* report*"; "decision model* report*"; "economic evaluation* report*"; "cost* evaluation* report*"; "simulation model* report*"; "multi modal comparison* report*"; "multimodal comparison* report*"; "infectious disease* practi*"; "communicable disease* practi*"; "transmission* model* practi*"; "decision* model* practi*"; "economic evaluation* practi*"; "cost evaluation* practi*"; "simulation model* practi*"; ""multimodal" comparison* practi*"; ""multi-modal" comparison* practi*"

#### 1.3. Inclusion/Exclusion Criteria

| **Inclusion** | **Exclusion** |
| --- | --- |
| - Reporting and best practice guidelines for dynamic models or decision-analytic models - Quality assessment tools for dynamic models or decision-analytic models | - Guidelines for clinical course models and drug or treatment assessments - Non-generalisable or irrelevant (to infectious disease or decision-analytic model) guidelines - Literature reviews unless they make suggestions of their own - Errata to, replies to, comments on, or summaries of existing articles - Conference abstracts - Individual cost-effectiveness or modelling studies - Articles presenting guidelines or recommendations which have subsequently been updated, for which the updated articles have been included - Elaboration and Explanation articles accompanying included articles - Non-English articles for which the English version has been included - Articles that cover only one single dimension |

### 2. Literature search for recent modelling studies

#### 2.1. Search results

| **Date of search for all databases: 22.01.2024** | |
| --- | --- |
| **Database/Register** | **Search** |
| **MEDLINE ALL (via Ovid)** 1946 to present | **9898** |
| **Total (after deduplication)** | **9854** |

#### 2.2. Search strategies

##### 2.2.1 MEDLINE ALL (via Ovid) (from 01.01.2019 to 22.01.2024)

1	Communicable Diseases/ep, pc, tm [Epidemiology, Prevention & Control, Transmission]

2	Disease Transmission, Infectious/pc [Prevention & Control]

3	(pandemic* or infectious disease* or epidemic* or communicable disease*).tw.

4	or/1-3

5	(peak? or (scenario? adj1 (future or projection*)) or outbreak or transmission?).tw.

6	(model or modelling or modeling or forecast or forecasts or forecasting or random forest* or nowcasting*).tw.

7	4 and 5and 6

8	limit 7 to yr="2019-Current"

#### 2.3. Inclusion

500 studies were randomly selected from the 9854 hits, and the 100 studies from these 500 that were deemed most relevant by a screener were included.
